# STUDY OF BLOOD GROUP ANALYSIS AND ITS CORRELATION WITH LYMPHOPENIA IN COVID-19 INFECTED CASES – OUR EXPERIENCE IN TERITARY CARE HOSPITAL

**DOI:** 10.1101/2021.07.12.21258824

**Authors:** Swarupa Ravuri, Saritha Cigiri, Harika Kalangi, Anunayi Jeshtadi, K.Nithesh Kumar, Nampally Vamshi Krishna, Srilatha Bacchu, K. Sudhakar

## Abstract

**AIMS AND OBJECTIVES:** To study the distribution and frequencies of ABO and Rhesus (Rh) blood groups among confirmed cases of Covid-19 infection. We also studied the relation between ABO blood group system and lymphopenia and studied the gender association in COVID-19 patients.

**METHODES:** A hospital based retrospective study was conducted at Government Medical College Suryapet from 01 Aug 2020 to 30 Sep 2020. A total of 200 Covid cases were included in the study who came to the hospital with the complaints of Fever, Sore throat, Body Pains, Cough, Breathlessness, Diarrhoea etc. Patients confirmed Covid infection was tested for blood grouping and RH typing by using forward blood grouping with the help of commercially available standard monoclonal antisera. CBP was processed in sysmax 5 part Haematology analyser. Blood group frequency was tested also assed the gender association, Covid patients presents with lymphopenia the relation between the ABO blood group and lymphocyte count was determined.

**RESULTS:** Males were more compared to the females .Middle aged group male patients were more commonly involved. Most predominant blood group was group B 79(39.5%), group O 78(39%),group A 37(18.5%), group AB 6(3%),most of them were 190 (95%)Rh positive, only 10 Rh negative (5%).To assess the Lymphopenia in our study we divided the absolute lymphocyte count into 5 groups. Group 1 cases are more 58 (29%), Group 2 91(45.5%), Group 3 30 (15%), Group 4 16(8%), Group 5 5(2.5%).

**CONCLUSION:** Male patients with blood group B were more compared to other blood groups however more number of studies are necessary to confirm these findings in a larger sample and among individuals of different ethnicities.

## INTRODUCTION

Corona virus disease also named Novel Corona Virus. First case was reported in December 2019 in Wuhan city and it was spread rapidly, more than 1,00,000 people were infected by the end of March 2020. This began in Wuhan wet-market, deals with animal trade business of different kinds of horse, bats, poultry, snakes, marmots. Recent reports have highlighted the role of zoonotic links, cross-species jumping and spill over conjuncture between animals and human transmission, before acquiring direct human-to-human contact. WHO announced a public health emergency of international concern. There are currently 7 known coronavirus that can infect human, Severe Acute Respiratory Syndrome (SARS), Middle East Respiratory Syndrome (MERS). Exact mode of transmission was unknown. Major routes of transmission person to person are through respiratory droplets and close contact with patients. Incubation period is 2-14 days, clinically patients with symptoms like fever, fatigue, dry cough and in severe cases presents with acute respiratory distress, septic shock, even death. Corona virus can affect the various systems in our body like Respiratory, Enteric, Neurological, Hepatic and renal.

## Pathogenesis

The mode of entry of this virulent pathogens is through respiratory droplets, tears, body fluids, mucous membranes of the eyes, mouth or nose & another mode of transmission through feco-oral route of transmission. Virus shows Tropism once it enters the body, type 2 pneumocystis and ciliated bronchial epithelial cells through angiotensin-converting enzyme 2 (ACE2) receptors and the immune cells like the dendritic cells and the macrophages. The incubation period is about 02 to 14 days post infection. By eliciting clinical data and details characteristics and determining, what were the risk factors may be associated with severity progression of the disease from the virus. Angiotensin converting enzyme 2 (ACE-2) host cell receptor plays a critical role in the entry of the virus into the cell to cause the infection. Variable expression of ACE-2 expression in the airway epithelial cells is one of the hypothesis for variable susceptibility to disease. In one study Gullion et al investigated the relation between natural antibodies of ABO blood system and ACE2 receptors, how these antibodies inhibit the adhesion of virus protein to the ACE receptors, he observed that S protein / ACE 2 dependent adhesion of Chinese hamster ovary cells to an ACE 2 expressing cell line could be inhibited by either monoclonal or anti-A antibodies may block the interaction between Corona virus and ACE 2 receptors providing the protection against the infection into the host. Blood tests have plays an important role in diagnosing the disease.

ABO blood grouping, discovered in 1900 by Austin immunologist Karl Landsteiner to explain the phenomena of red blood cell (RBC) agglutination, is well-documented hypothesis. Landsteiner carbohydrate moieties are genetically inherited. These moieties suggest correlation between ABO blood type, cardiovascular disease, cancer and certain infections including SARS virus. In humans 4 types of ABO blood groups namely A, AB, B, O, located on chromosome 9 (9q34.2). Blood group antigens are one of the main antigens named human histo-blood group antigens (HBG’S) located on surface of RBC’S. Difference in the expression of antigen can increase or decrease host susceptibility to many infections. Blood groups O have significantly high risk of hepatitis B. HBV infection more in Rh-Positive Donors. Rotavirus gastro enteritis was significantly more prevalent among blood group A patients, malaria patients with blood group A had high risk of anaemia compared to another blood groups. AB blood group had high risk of developing Dengue haemorrhagic fever O blood group patients more susceptible to norovirus infection. SAR’S Corona virus is a new virus we have to find out which blood group individuals susceptible to Covid 19 infection. A retrospective study was conducted to explore the relationship and also lymphopenia is a common feature of Covid 19 patients associated with severity disease. The association between ABO blood groups and lymphocytes count was also investigated.

## Materials & Methods

Materials & Methods:
|  |  |
| --- | --- |
| **Study design:** | Descriptive, Retrospective & prospective study |
| **Study population:** | Patients who referred to department of pathology |
| **Location of study:** | Government Medical College/Hospital, Suryapet, Telangana, India |
| **Study duration:** | 2 Months |
| **Inclusion criteria:** | Patients with above 18 years were included |
| **Exclusion criteria:** | Patients with below 18 years were excluded |
| **Sampling method:** | Consecutive sampling method |
| **Sample size:** | A total of 200 were included |

### METHODES

A retrospective study was performed at Government Medical College/ Hospital Suryapet, Telangana. In this study 200 patients who have admitted in the hospital with the symptoms of Covid 19 at least one symptom or sign of cough, respiratory distress, fever or features that are unexplained by any other disease, history of travel to other country, contact with the relatives or close contact with the patients that who are confirmed positive for Covid 19 by Real Time PCR (RT-PCR) in the last 14 days. Patients with any of the above said symptoms of Covid 19 comes to us during the study period from 01 Aug 2020 to 30 Sep 2020 were enrolled in Government Medical College Suryapet, all were subjected to demographic, clinical feature, laboratory findings. Demographics include age, gender, hypertension, diabetics and heart diseases. Laboratory investigations include Throat swab test and RT-PCR confirmed cases were further investigated CBP, NLR, PLR, blood grouping and RH typing, LFT, Creatinine, LDH. Only the results of RT-PCR, CBP, blood grouping were used for this study. CBP was processed in Sysmax 5 part analyser, and blood grouping was performed with standard protocols. Blood grouping was determined by forward grouping (cell grouping) by agglutination methods with the help of commercially available standard monoclonal antisera A, antisera B, antisera AB and antisera D manufactured by Tulip Diagnostics (p) ltd Verna Goa. All the blood group O were tested by anti-H lectin Tulip Diagnostics (p) ltd Verna Goa to rule out Bombay blood group. These antisera mixed with patient blood and check for agglutination accordingly blood group is determined. Rh typing was done to see the Rh factor on the surface of the red blood cells. Rh positive if cells have this surface protein, Rh Negative if you do not have this cell surface protein. All information was obtained and analysed with standard excel programme.

### ASSOCIATION ANALYSIS

The association between different blood types and Covid 19. According to gender, subgroups were stratified to assess whether there was a significant difference between blood type and the incidence of COVID 19. Correlation analysis between blood group and lymphocytes count was also performed.

### STATISTICAL ANALYSIS

Statistical analysis was carried out using the Statistical Package for Social Sciences (SPSS) VERSION 21.0. Independent sample t-tests were used for age, white blood cell count, lymphocyte count, neutrophil lymphocyte ratio. Chi-square test with 95% confidence intervel is used to test ABO blood group frequency in all populations and different gender subgroup was tested. Analysis association between the ABO blood grouping and the lymphocyte count was performed with analysis of variance (ANOVA) AND A LINEAR REGRESSION MODEL. A p<0.05 was considered significant.

## RESULTS

A total of 200 patients were included in this study where Males were 135 (67.5%) & females were 65 (32.5%). Age distribution was shown in the table No.01. Most of the patients belong to 50 years (25.5%) of age group & 60 years (21%). Maximum age was 85 years, minimum age 19 years and mean age was 50 years.

**Table No.01.**
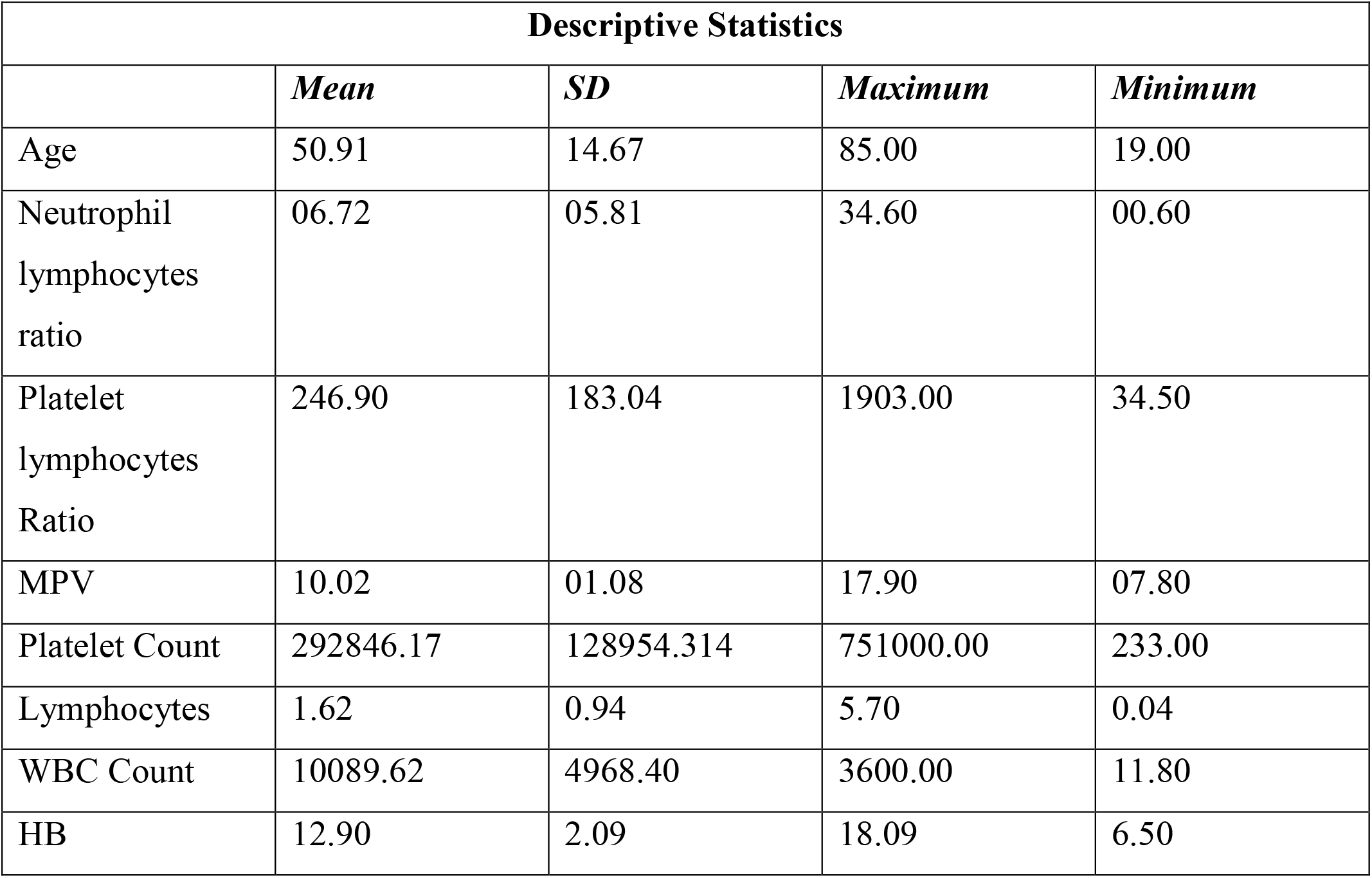
shows the age distribution of patients.

STUDY OF BLOOD GROUP ANALYSIS AND ITS CORRELATION WITH LYMPHOPENIA IN COVID-19 INFECTED CASES – OUR EXPERIENCE IN TERTIARY CARE HOSPITAL
| Descriptive Statistics |  |  |  |  |
| --- | --- | --- | --- | --- |
|  | *Mean* | *SD* | *Maximum* | *Minimum* |
| Age | 50.91 | 14.67 | 85.00 | 19.00 |
| Neutrophil lymphocytes ratio | 06.72 | 05.81 | 34.60 | 00.60 |
| Platelet lymphocytes Ratio | 246.90 | 183.04 | 1903.00 | 34.50 |
| MPV | 10.02 | 01.08 | 17.90 | 07.80 |
| Platelet Count | 292846.17 | 128954.314 | 751000.00 | 233.00 |
| Lymphocytes | 1.62 | 0.94 | 5.70 | 0.04 |
| WBC Count | 10089.62 | 4968.40 | 3600.00 | 11.80 |
| HB | 12.90 | 2.09 | 18.09 | 6.50 |

Blood group distribution shown in the PIE diagram No. 01

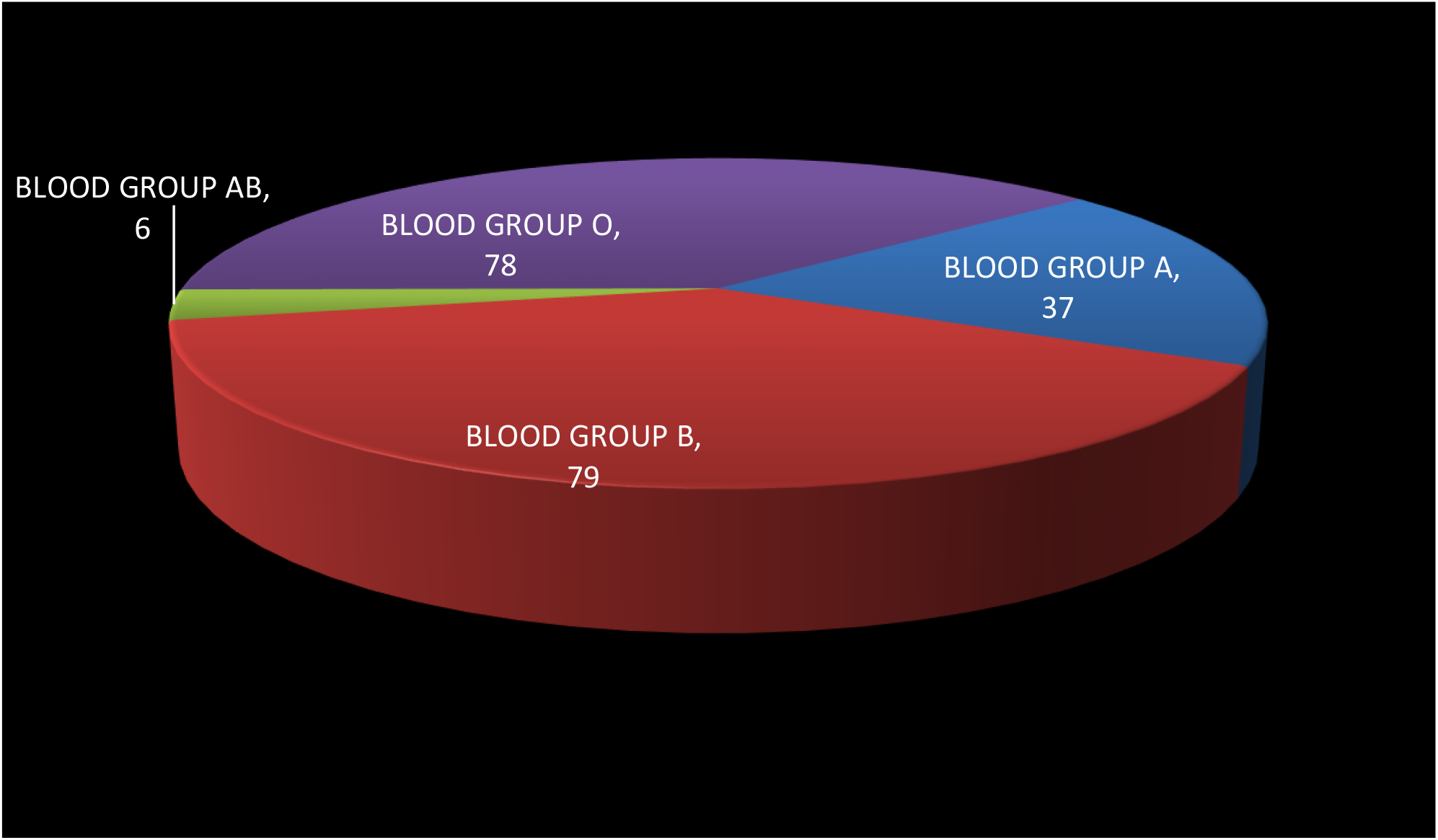

Most predominant blood group was group B 79(39.5%), group O 78(39%),group A 37(18.5%), group AB 6 (3%),most of them were Rh positive only 10 Rh negative (5%).

In addition lymphocyte count depletion is a related to severity of the disease we also studied the relation between lymphopenia and blood group association shown in Table 2

**Table No.2.**
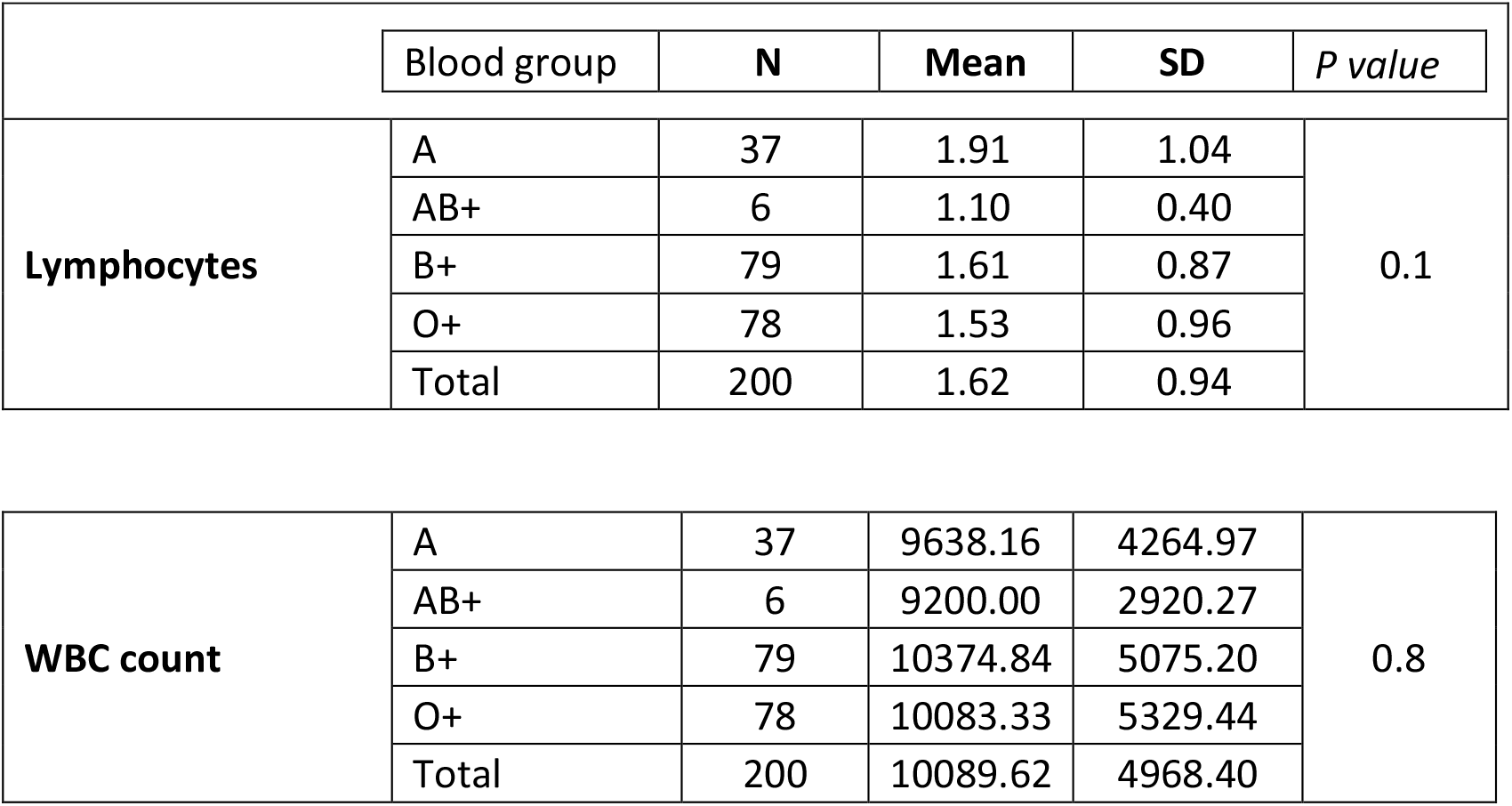
showing the lymphocyte count and WBC count in association between the different blood groups.

**Table 3.**
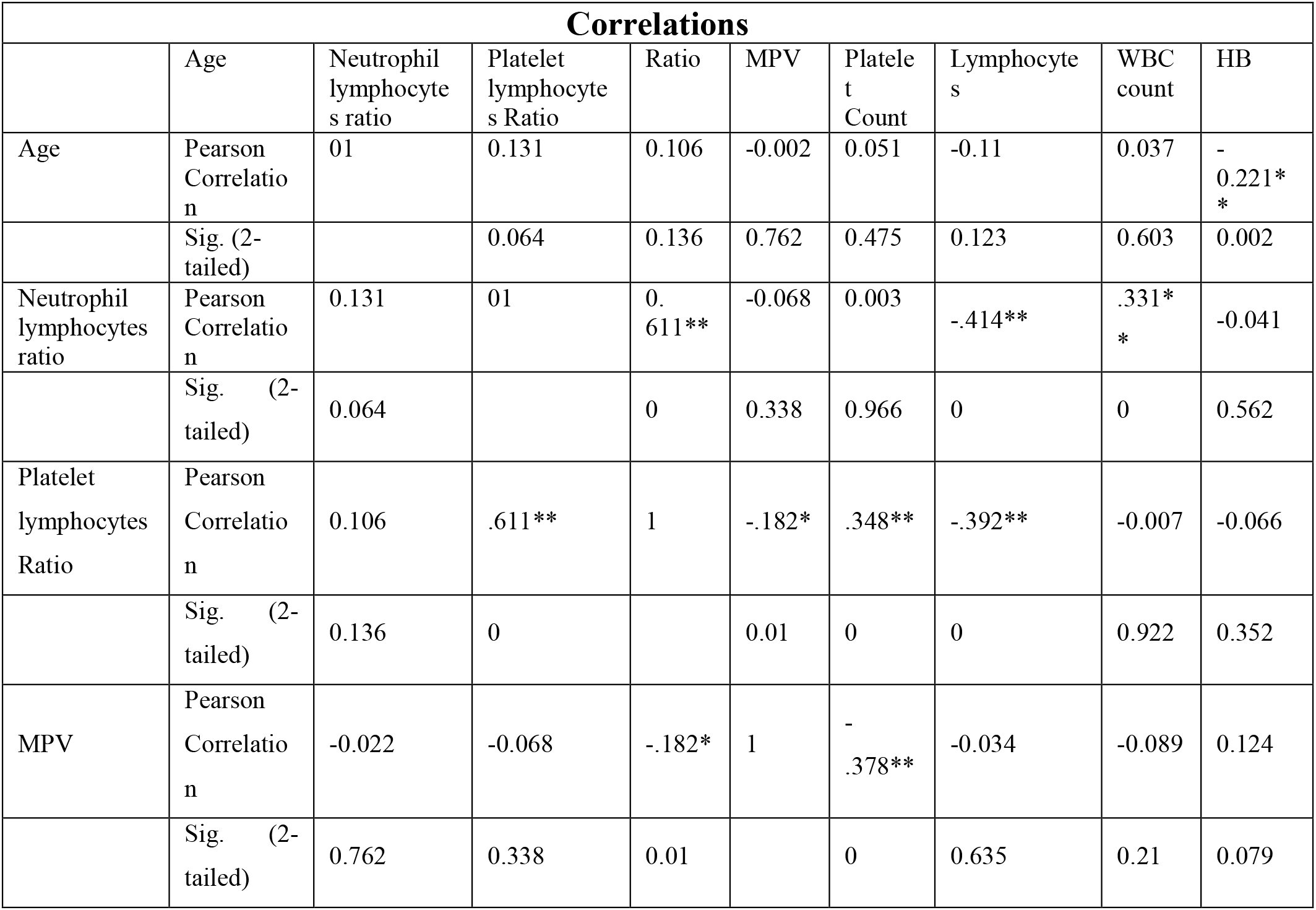
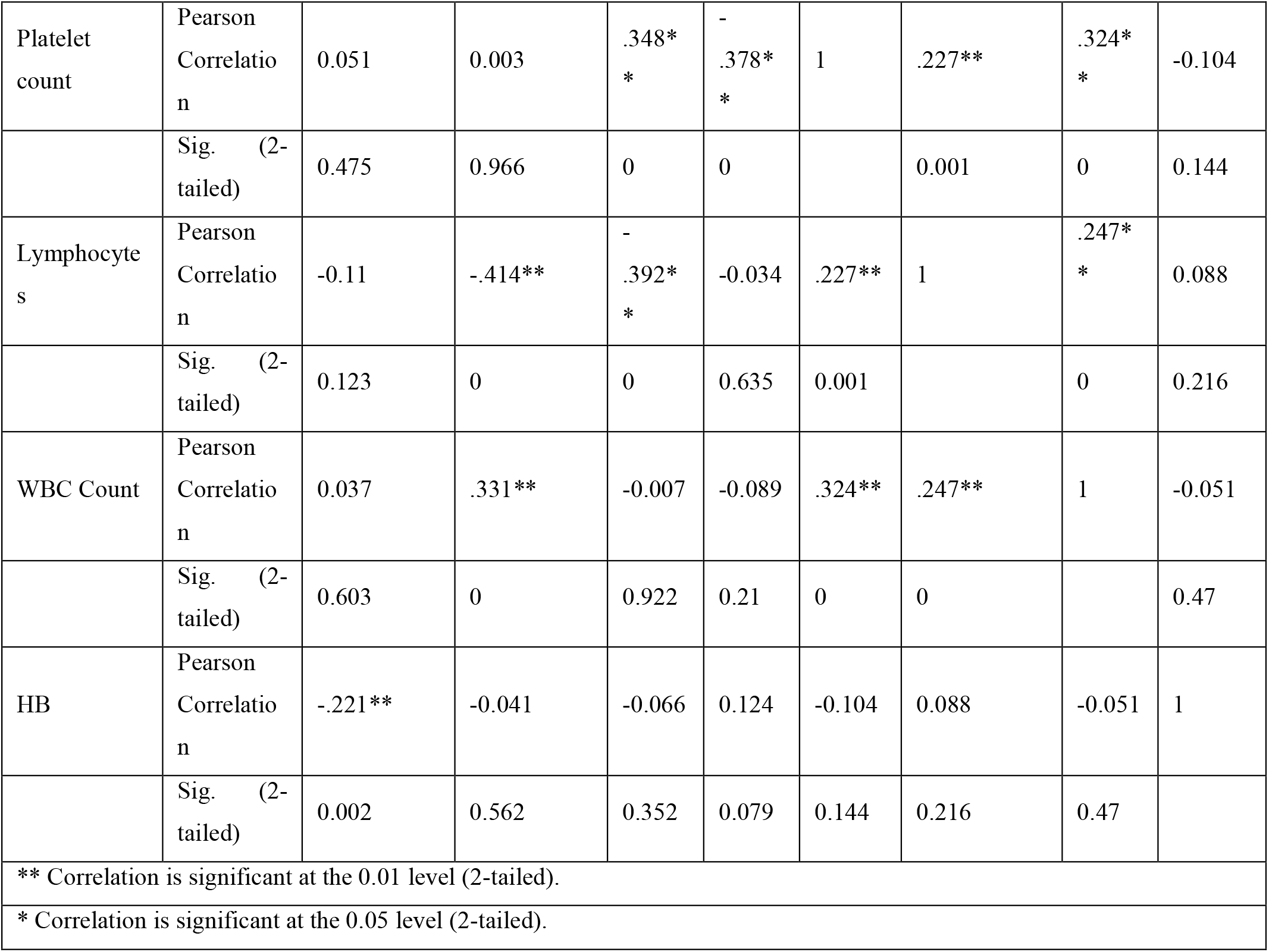
shows Pearson correlation between different parameters and values less than 0.01 found to be significant.

Lymphopenia in our study we divided the absolute lymphocyte count into 5 groups. Group1 0- 1.0 Group 2 1.1 – 2.0, Group 3 2.1-3.0, Group 4 3.1-4.0, Group 5 >4

Group 1 cases are more 58 (29%), Group2 91(45.5%), Group 3 30 (15%), Group 4 16(8%), Group 5 5(2.5%). Mean lymphocyte count was 1.62 minimum was 0.04 and maximum was 5.70. P-value counted in different blood groups it was 0.1 found to be significant. Association between lymphocyte count WBC count and blood groups had shown in box diagrams 01& 02.

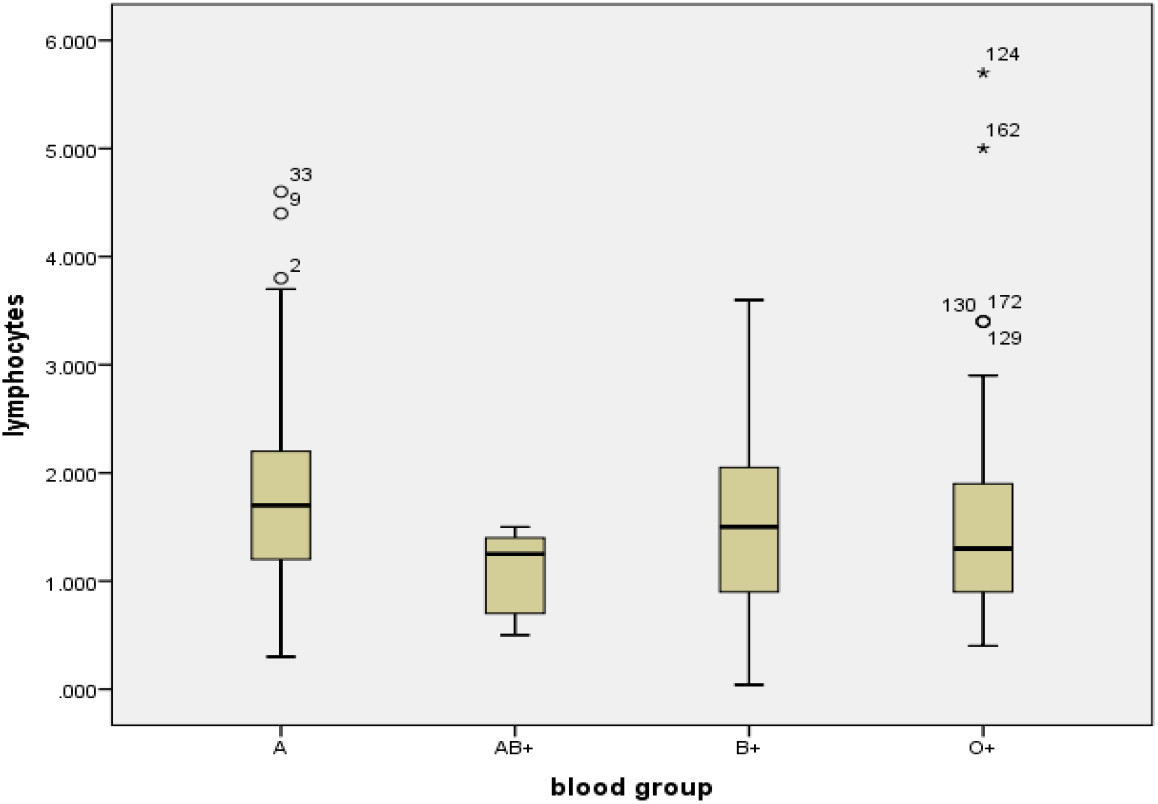
Box digram 1 showing the association between blood grouping and lymhocyte count.

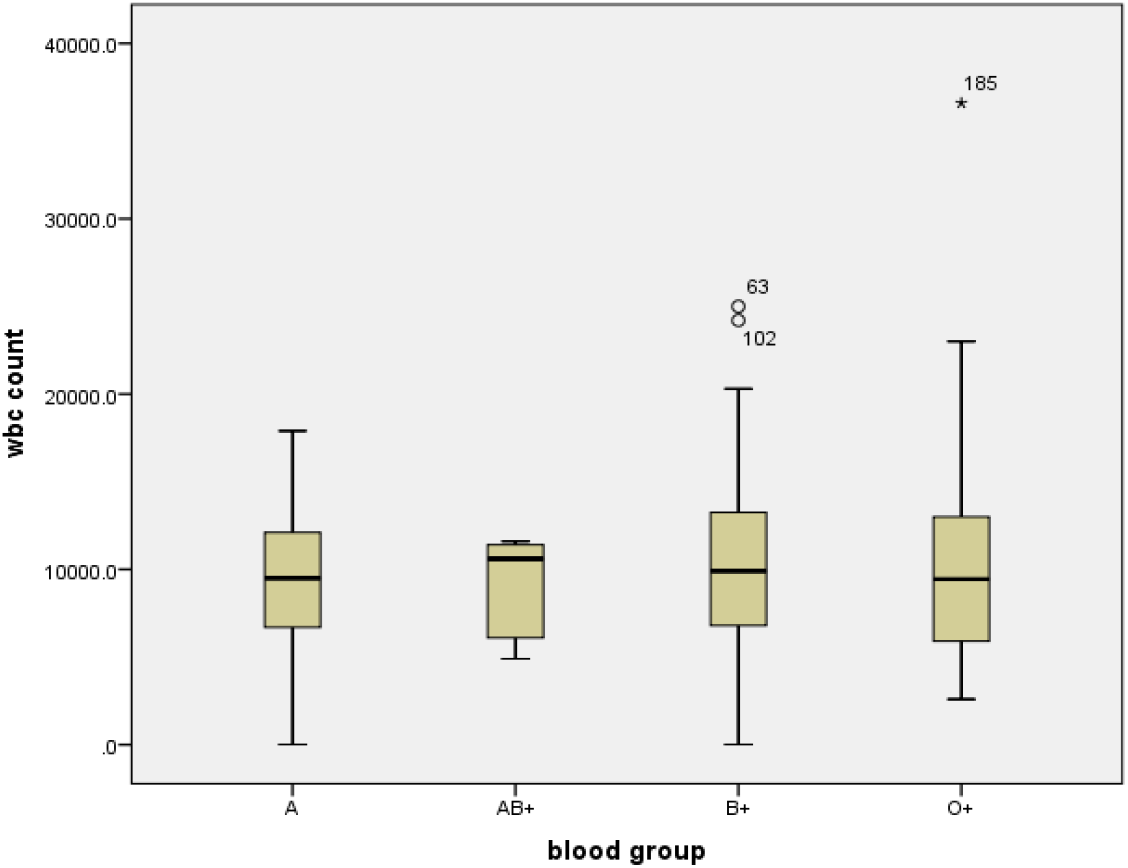
Box diagram 2 showing the association between blood grouping and WBC count

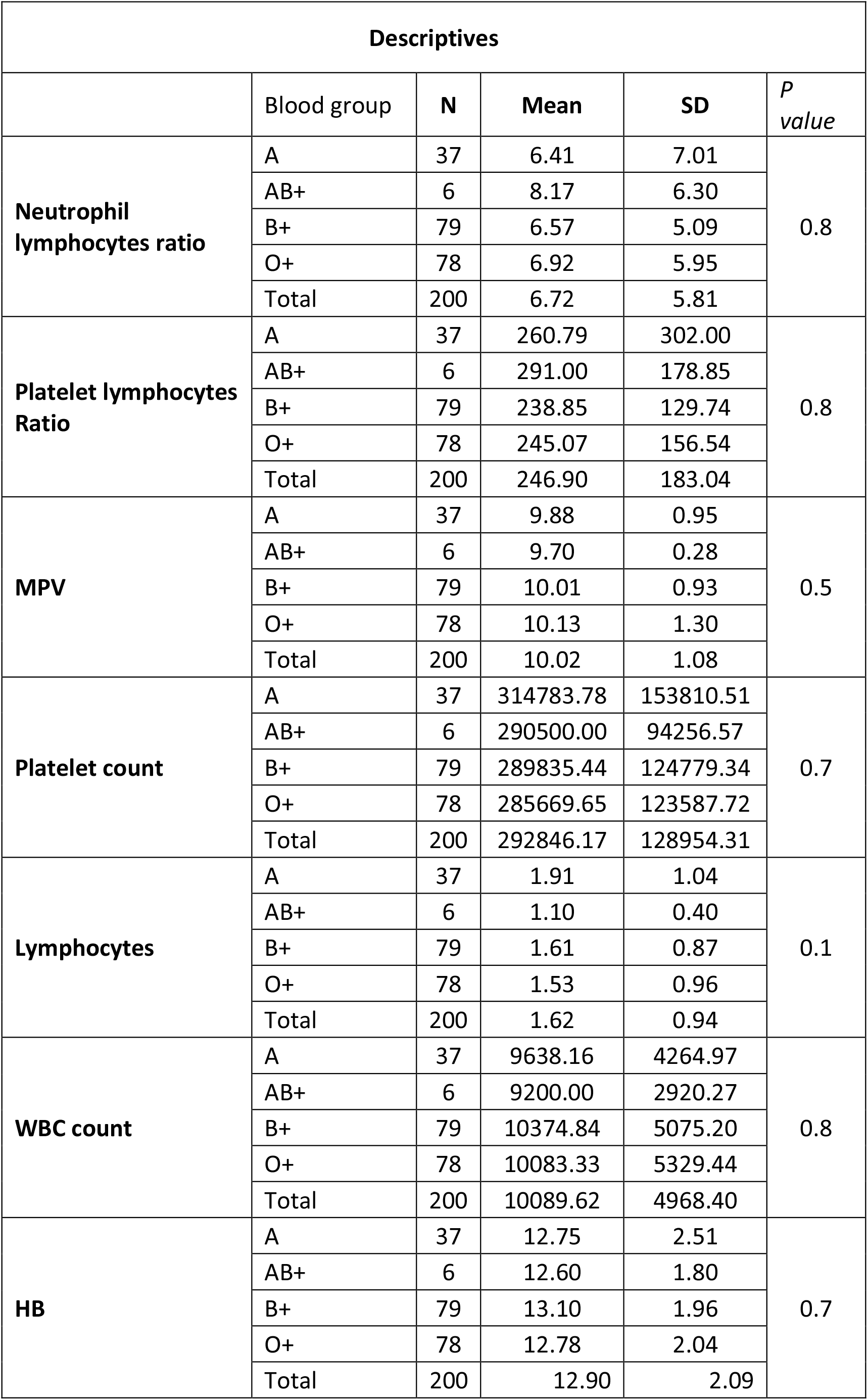
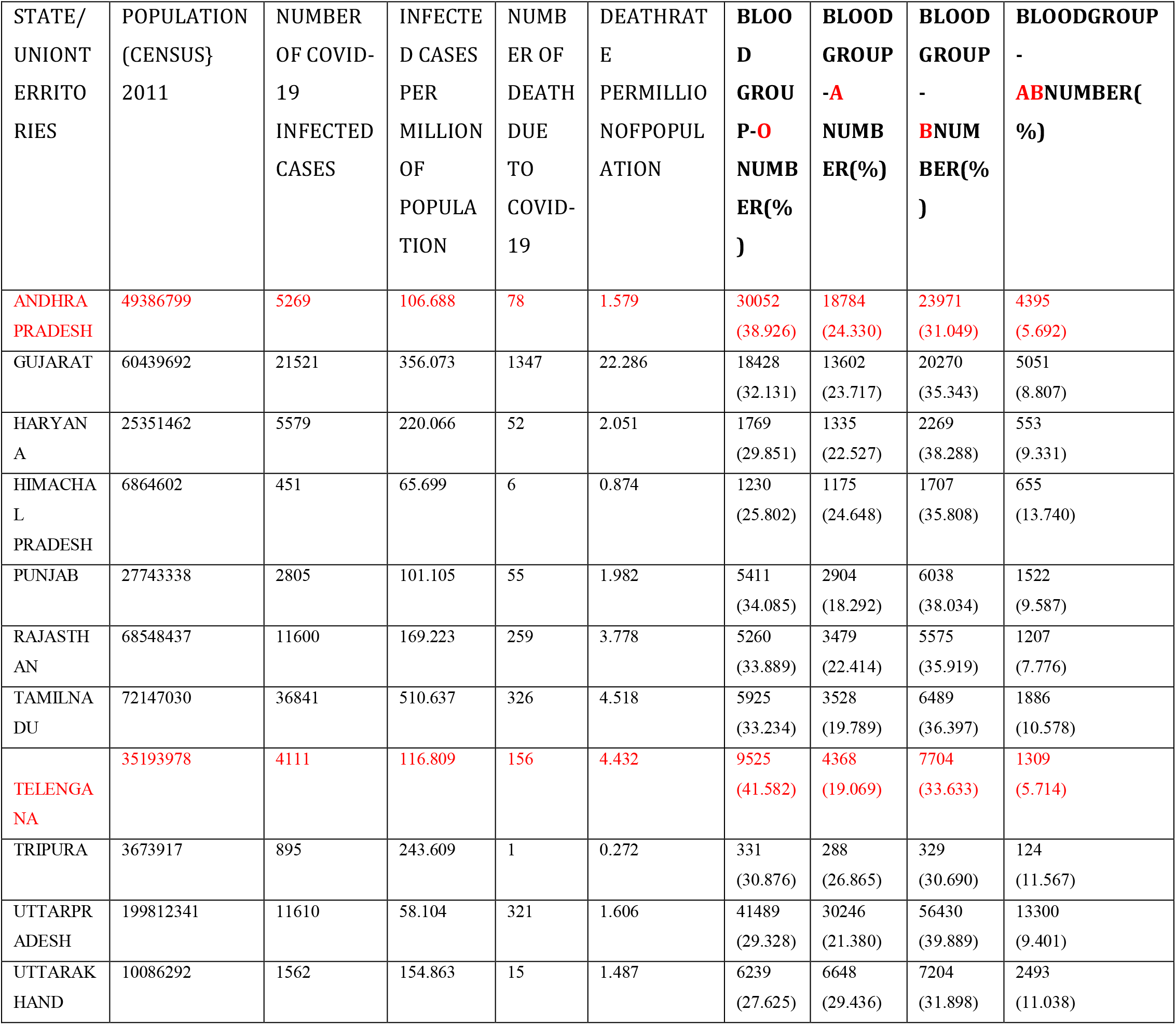

Studies from other counties Li et al, Zhao et al studies they found that the blood group A patients infected with SARS CoV 2 higher than the healthy controls and blood group O in SARS CoV 2 was low, our study not correlating with these two studies, another study conducted by Michel zietz study blood group B association with covid19 infection is more than the other blood groups.

Zhao et al 2020 conducted a study with 2173 patients with covid confirmed patients it was meta-analysis study they concluded that patient’s ABO blood group and the patients with blood group A were associated with high risk when compared to other non A blood groups, O blood group was associated with lower risk for the infection compared to others.

Yamamoto et al 2020 study concluded that s protein produced in A, AB, B blood group individuals with their antigens present on the surface of the red blood cells would produce corresponding respective antibodies. These antibodies would block the interaction with S-protein and ACE receptors.

Tanigawa et al in 2020 reviewed the current literature on COVID19 and associated phenotypes. It was a study conducted in UK bio bank with 337579 cases, in this study they aggregated human leucocyte antigen and ABO blood type frequencies. They found that blood group O show consistent risk reduction in accordance with Zhao et al study. The mechanism of association and susceptibility of patients with Blood group B among patients it is unknown. Normally interaction between the spike proteins andACE-2 receptors is hampered by the antiblood type A, which gives the explanation of protection of blood group O against the COVID 19 infection and mortality. The role of ABO antibodies altering the interaction between the spike proteins of SARS Cov 2 and ACE 2 RECEPTORS still unknown. For the multiplication of virus and spread of infection to another hosts, the spike protein of virion carries A, B, AB glycan antigen depends on the blood group of carrier. Blood group O patients contains both A & B antibodies, infection rate with A, B & AB antigens would be reduced. Protection also depends on the ABO antibody titers. These variation of observations on association of blood groups and COVID 19 infection could be due to different strains of SARS Cov 2 with varying pathogenicity and discrepancies in in the investigated population.

In our research we also studied the relationship between ABO blood grouping & lymphocytes Count. We also further investigated the importance of Lymphocyte count in the evaluation of severity of Covid 19 infection. The lymphocyte count in B type blood group patients mean was 1.91. AB blood group patients were 6 in number this blood group patients present with low lymphocyte count 1.10 with high NLR values and P-value calculated 0.1 found to be significant.

Preventive measures such as social distancing, hand hygiene, home or hospital quarantine as well as restrictions on movement (lockdown) help in preventing the spread of the disease. Isolation and supportive therapy, including ventilator support, are the mainstays of treatment of infected patients.

### Limitations

Meanwhile several drawbacks existed in our study, limited sample size of Covid – 19 patients, second regional selection bias. This is a observational study attempts were made to control for confounding.

## Conclusion

Male patients with blood group B were more effective when compared to other blood groups, however more number of studies are necessary to confirm these findings in a larger sample and among individuals of different ethnicities.

## Data Availability

No

---

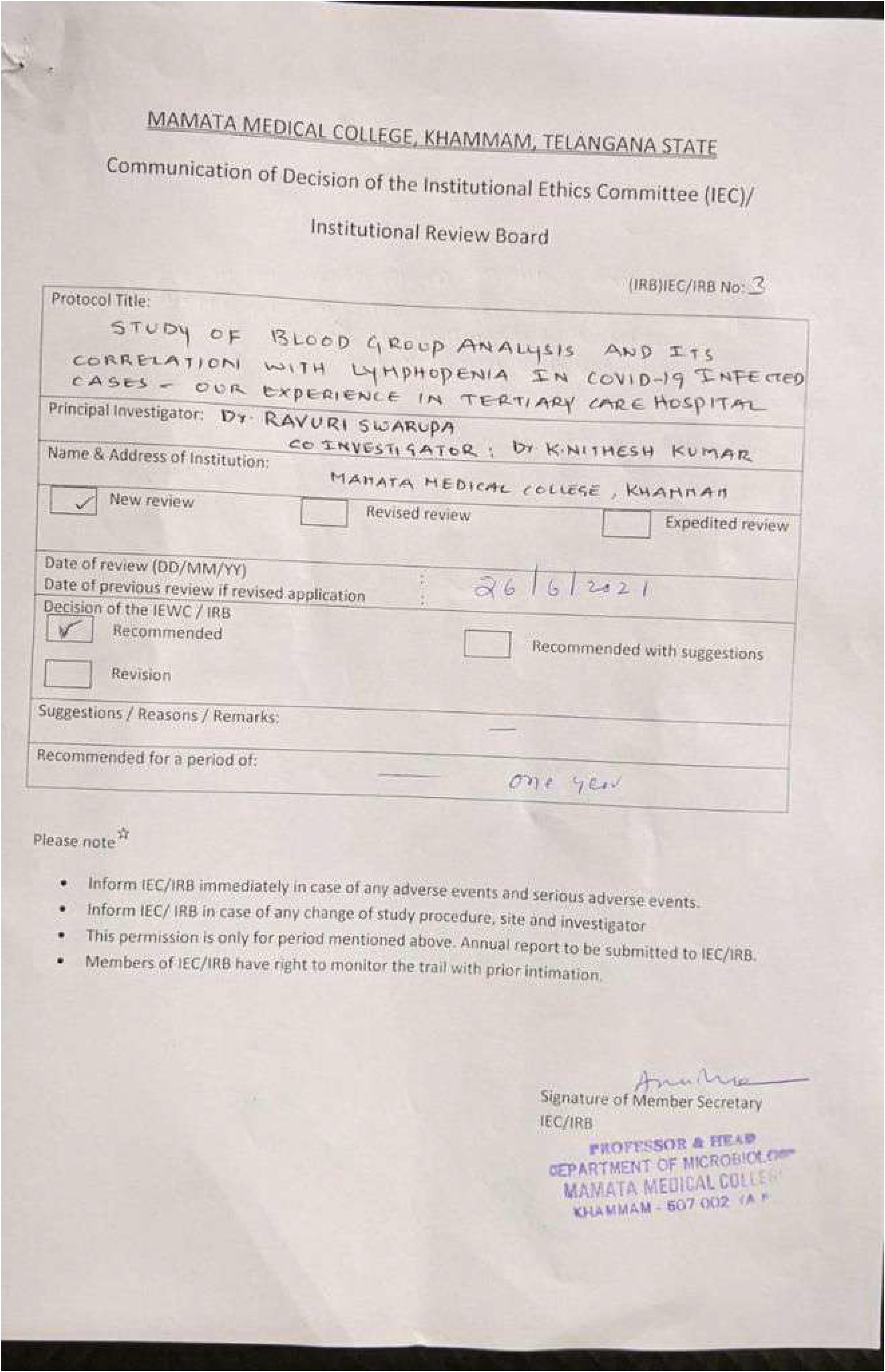

## BIBILOGRAPHY

1. Wang, D., Hu, B., Hu, C., Zhu, F., Liu, X., Zhang, J., et al.(2020). Clinical Characteristics of 138 Hospitalized Patients With 2019 Novel Coronavirus-Infected Pneumonia in Wuhan, China. JAMA. 323, 1061–1069. doi: 10.1001/jama.2020.1585 Wang, D., Hu, B., Hu, C., Zhu, F., Liu, X., Zhang, J., et al. (2020). Clinical Characteristics of 138 Hospitalized Patients With 2019 Novel Coronavirus-Infected Pneumonia inWuhan, China. JAMA. 323, 1061–1069. doi: 10.1001/jama.2020.1585.

2. Wu, Z., and McGoogan, J. M. (2020). Characteristics of and Important LessonsFromthe CoronavirusDisease 2019 (COVID-19)Outbreak in China: Summaryof a Report of 72314 Cases From the Chinese Center for Disease Control andPrevention. JAMA. 323, 1239–1242. doi: 10.1001/jama.2020.2648.

3. Huang, C.,Wang, Y., Li, X., Ren, L., Zhao, J., Hu, Y., et al. (2020). Clinical featuresof patients infected with 2019 novel coronavirus inWuhan, China. Lancet 395,497–506. doi: 10.1016/S0140-6736(20)30183-5.

4 Bennett J,Dolin R, Blaser MJ. Principles and Practicr of infectious Disease. 8th Edition Elsevier / Saunders, PA, usa(2014).

5. Meng J, Xiao G, Zhang J, He X, Ou M, Bi J, Yang R, di W, Wang Z, Li Z, Gao H, Liu L, Zhang G (2020) Renin-angiotensin systeminhibitors improve the clinical outcomes of COVID-19 patients with hypertension. Emerg Microbes Infect 9(1):757–760.

6. Hussain A, Bhowmik B, do Vale Moreira NC (2020) COVID-19and diabetes: knowledge in progress. Diabetes Res Clin Pract 162:108142.

7. GuoW, Li M, Dong Y, et al (2020) Diabetes is a risk factor for theprogression and prognosis of COVID-19 [published online aheadof print, 2020 Mar 31]. DiabetesMetab Res Rev. https://doi.org/10.1002/dmrr.3319.

8. Zhao Q,Meng M, Kumar R, et al (2020) The impact of COPD andsmoking history on the severity of COVID-19: A systemic reviewand meta-analysis. J Med Virol. https://doi.org/10.1002/jmv.25889.

9 Wrapp, D., Wang, N., Corbett, K. S., Goldsmith, J. A., Hsieh, C. L., Abiona, O., et al. (2020). Cryo-EM structure of the 2019-nCoV spike in the prefusion conformation. Science 367, 1260–1263. doi: 10.1126/science.abb2507

10 Leung JM, Yang CX, Tam A, Shaipanich T, Hackett TL, Singhera GK, Dorscheid DR, Sin DD (2020) ACE-2 expression in the smallairway epithelia of smokers and COPD patients: implications forCOVID-19. Eur Respir J 55:2000688.

11. Guillon, P., Clement, M., Sebille, V., Rivain, J. G., Chou, C. F., Ruvoen-Clouet, N.,et al. (2008). Inhibition of the interaction between the SARS-CoV spike proteinand its cellular receptor by anti-histo-blood group antibodies. Glycobiology 18,1085–1093. doi: 10.1093/glycob/cwn093

12 Melzer, D., Perry, J. R., Hernandez, D., Corsi, A. M., Stevens, K., Rafferty, I., et al. (2008). A genome-wide association study identifies protein quantitative trait loci (pQTLs). PLoS Genet. 4:e1000072. doi: 10.1371/journal.pgen.1000072.

13. Wiggins, K. L., Smith, N. L., Glazer, N. L., Rosendaal, F. R., Heckbert, S. R., Psaty, B.M., et al. (2009). ABO genotype and risk of thrombotic events and hemorrhagicstroke. J. Thromb. Haemost. 7, 263–269. doi: 10.1111/j.1538-7836.2008.03243.

14. Mohammadali, F., and Pourfathollah, A. (2014). Association of ABO and Rh bloodgroups to blood-borne infections among blood donors in Tehran-Iran. Iran. J.Public Health 43, 981–989.

15. Elnady, H. G., Abdel, S. O., Saleh, M. T., Sherif, L. S., Abdalmoneam, N., Kholoussi, N. M., et al. (2017). ABO blood grouping in Egyptian children with rotavirus gastroenteritis. Prz. Gastroenterol. 12, 175–180. doi: 10.5114/pg.2017.70469.

16. Murugananthan, K., Subramaniyam, S., Kumanan, T., Owens, L., Ketheesan, N., and Noordeen, F. (2018). Blood group AB is associated with severe forms of dengue virus infection. Virusdisease 29, 103–105.doi: 10.1007/s13337-018-0426-8.

17. ABObloodgroupsystemisassociatedwithCOVID19mortality:AnepidemiologicalinvestigationintheIndianpopulationSunali Padhi1,SubhamSuvankar1,DebabrataDash,VenketeshK.Panda,AbhijitPati,JogeswarPanigrahi,AdityaK.Panda*Departmentof BioscienceandBioinformatics,KhallikoteUniversity,TransitCampus:GMaxBuilding,Konisi,Berhampur,761008Odisha,India. https://doi.org/10.1016/j.tracli.2020.08.00912467820/©2020Soci’et’efrancąisedetransfusionsanguine(SFTS). PublishedbyElsevierMassonSAS.Allrightsreserved.transfusion Cliniqueetbiologicue27(2020)253–258.

18. Li, J., Wang, X., Chen, J., Cai, Y., Deng, A., and Yang, M. (2020). Associationbetween ABO blood groups and risk of SARS-CoV-2 pneumonia. Br. J.Haematol. 190, 24–27. doi: 10.1111/bjh.16797.

19 Zhao, J., Yang, Y., Huang, H., Li, D., and Gu, D., Lu, X., et al. (2020). Relationshipbetween the ABO blood group and the COVID-19 susceptibility. MedRxiv.doi: 10.1101/2020.03.11.20031096. [Epub ahead of print].

20. Yamamoto F. ABO Blood Groups and SARS-CoV-2 Infection; 2020.

21. Saxena S. Coronavirus disease-2019: A brief compilation of facts. J Oral Maxillofac Pathol 2020;24:5–7.

22. Breiman A, Ruvën-Clouet N, LePendu J. Harnessingthenaturalanti-glycanimmuneresponse to limit the transmission of enveloped viruses suchasSARS-CoV-2. PLoSPathog 2020;16:e1008556.

